# Linking Network Damage to Post-Stroke Depression: A Neurotransmitter-Informed Connectome Analysis

**DOI:** 10.64898/2026.04.24.26351561

**Authors:** Benedikt M. Frey, Julian Klingbeil, Margaret Jane Moore, Philipp J. Koch, Jan Feldheim, Tom Hornberger, Götz Thomalla, Tim Magnus, Fanny Quandt, Nele Demeyere, Dorothee Saur, Robert Schulz

**Affiliations:** Department of Neurology, University Medical Center Hamburg-Eppendorf, Germany; Department of Neurology, University Medicine Halle, Halle, Germany; Department of Neurology, University of Leipzig Medical Center, Germany; Queensland Brain Institute, University of Queensland, Brisbane, Australia; Department of Neurology, Charité Berlin, Germany; Department of Experimental Psychology, Radcliffe Observatory Quarter, University of Oxford, Oxford, United Kingdom

## Abstract

Post-stroke depressive symptoms (PSDS) are a frequent and disabling consequence of stroke. While lesion-network studies implicate disruption of large-scale affective circuits in PSDS, the neurobiological factors determining why certain network disruptions confer vulnerability to PSDS remain insufficiently understood. We analyzed data from two independent stroke cohorts (total n = 435). Acute lesion masks were embedded within normative structural connectomes, weighted by positron-emission tomography-derived maps of 19 neurotransmitter receptors and transporters, to quantify neurotransmitter (NT)-informed network damage. Partial least squares regression with variable importance measures was used to identify NT-specific damage scores that were informative for PSDS, as quantified by the Hospital Anxiety and Depression Scale at follow-up. Informative NT-systems were subsequently evaluated in multivariable logistic regression models adjusted for age, sex, lesion volume, and neurological deficit. Across cohorts, multivariate analyses converged on a neurochemical signature involving serotonergic, cholinergic, dopaminergic, and GABAergic networks. Damage to networks related to the serotonin transporter (5-HTT) and the vesicular acetylcholine transporter (VAChT) was independently associated with increased odds of PSDS in covariate-adjusted models and improved model fit beyond clinical and lesion-based predictors. In contrast, associations with other NT systems, including dopaminergic networks, were not consistently implicated across cohorts. These findings identify the serotonergic and cholinergic network architecture as a key neurochemical substrate that modulates vulnerability to PSDS. By integrating structural disconnection mapping with NT-informed connectomics, this study provides a mechanistic framework that links stroke-induced network disruption to PSDS and highlights serotonergic and cholinergic systems as central pathways for hypothesis-driven risk stratification and future multimodal investigations.

## Introduction

Post-stroke depressive symptoms (PSDS) affect approximately one-third of stroke survivors and represent one of the most frequent and disabling neuropsychiatric sequelae of cerebrovascular disease ^1^. PSDS are associated with poorer functional recovery, reduced quality of life, increased dependency, higher levels of cognitive impairment, and increased long-term mortality ^2-5^. A broad set of clinical, psychosocial, and demographic factors, including stroke severity, disability, premorbid vulnerability, cognitive impairment, and reduced social support, contribute to risk, yet together do not fully explain the substantial interindividual variability in PSDS ^6^.

Historically, attempts to localize depression to focal lesion sites have yielded inconsistent results. Early influential reports proposed links between post-stroke depression and left frontal lesions, while also noting that many depressed patients had lesions outside the left frontal cortex ^41,42^. This result has failed to replicate or exhibited time-dependency, with multiple systematic reviews and meta-analyses reporting no reliable relationship between specific lesion locations, particularly left frontal lesions, and depression after stroke ^2,43-45^. Likewise, voxel-based lesion-symptom mapping studies have generally not identified consistent lesion loci for depression ^34,46,47^. These observations align with the broader methodological insight that symptoms may not map to single regions, but rather to distributed circuits and their remote effects (diaschisis) ^39,48^. Accordingly, neuroimaging research has increasingly conceptualized post-stroke depression as a network disorder, linking depressive symptoms to dysfunction within distributed fronto-limbic and fronto-striatal circuits rather than to focal lesion locations alone ^6,8,34,49,50^. Lesion network mapping has provided a principled explanation for why heterogeneous lesion locations may converge on a common circuit, showing that lesions associated with depression can be unified by their functional connectivity to a left dorsolateral prefrontal circuit and that “network damage” can predict depression prevalence and severity ^7^. Studies using disconnection-based prediction further reinforce the notion that structural network disruption is related to PSDS, yet the reported networks and hubs still vary across cohorts. Rather than implying the absence of a consistent lesion-depression relationship, this variability likely reflects differences in lesion distributions, clinical characteristics, and statistical power across samples, even in the presence of a shared underlying network vulnerability. Importantly, network-based lesion models explain only a limited proportion of variance in depressive symptoms, and a substantial subset of patients develop PSD despite preserved integrity of proposed critical regions or circuits. This highlights that lesion and network damage, while informative, are insufficient to fully account for individual risk, motivating the investigation of complementary predictors that may capture additional biological or clinical dimensions relevant for depression after stroke ^8,9^.

At the molecular level, monoaminergic systems play central roles in mood regulation, motivation, and emotional learning ^10^. In particular, serotonergic mechanisms are implicated in major depressive disorder and PSDS, supported by genetic findings and pharmacological evidence with SSRIs ^6,11-13^. However, traditional lesion and network approaches generally do not specify which molecular architectures render certain networks vulnerable. Recent work has therefore moved toward biologically annotated connectomics, integrating PET-derived receptor/transporter maps with network models to link macro-scale disconnection to neurochemically constrained network architecture ^14-16^.

In the present study, we apply this integrative framework to identify neurotransmitter systems whose spatial organization confers selective vulnerability to PSDS. By combining neurotransmitter-informed connectomics with multivariate predictor selection and covariate-adjusted outcome modeling across two independent cohorts, we aim to develop a mechanistic model that integrates stroke-induced network disruption with molecular brain architecture. We hypothesized that PSDS would be associated not only with stroke-related disability, stroke lesion volume, and structural disconnection, but also that NT-informed network disruption, particularly within serotonergic and dopaminergic systems, would add explanatory value beyond established models.

## Methods and Materials

### Participants

This study reanalyzed published neuropsychological and imaging data from 435 stroke patients across two independent datasets, Leipzig ^17^ and Oxford ^18,19^. While the Leipzig dataset was derived from a monocenter study, the Oxford dataset included data from two cohorts: the OCS-Care and OCS-Recovery studies. Details on inclusion criteria and patient recruitment can be found in the original papers. The primary outcome of this study was the presence of PSDS, as defined as a score >7 in the depression subscale of the Hospital Anxiety and Depression Scale (HADS-D) administered six months post-stroke. Key control variables were age, sex, stroke lesion volume, and stroke-related disability, as quantified by the National Institutes of Health Stroke Scale (NIHSS) and the Barthel Index (BI). A total of 601 unique stroke patients were included (Leipzig n = 270; Oxford n = 331). After excluding participants with missing key clinical variables and only grey matter lesions, the final sample for subsequent statistical analyses comprised 435 cases with complete data on age, sex, stroke-related disability, and PSDS at follow-up, including 267 from the Leipzig and 168 from the Oxford datasets. Table 1 summarizes demographic, clinical, and imaging data. Local institutional review boards approved all original study protocols. All procedures were in accordance with the Declaration of Helsinki.

**Table 1.**
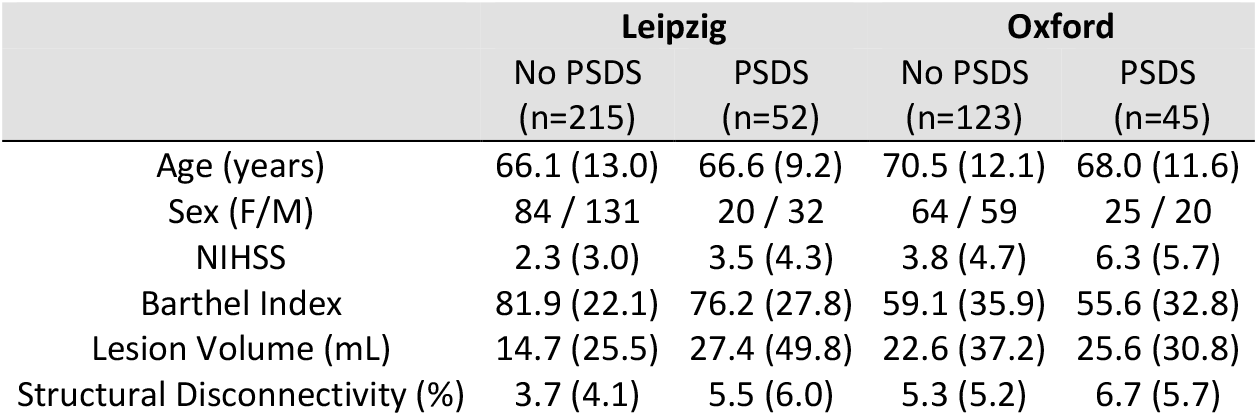
Demographic, clinical data, and imaging data of the included participants. Demographic and clinical characteristics of both cohorts. Values are given in mean (SD) and total number, respectively. Disconnection severity is reported as a percentage of 2 million normative streamlines. PSDS: post-stroke depressive symptoms, defined as a HADS-D depression score >7 at follow-up.

|  | Leipzig |  | Oxford |  |
| --- | --- | --- | --- | --- |
|  | No PSDS<br>(n=215) | PSDS<br>(n=52) | No PSDS<br>(n=123) | PSDS<br>(n=45) |
| Age (years) | 66.1 (13.0) | 66.6 (9.2) | 70.5 (12.1) | 68.0 (11.6) |
| Sex (F/M) | 84 / 131 | 20 / 32 | 64 / 59 | 25 / 20 |
| NIHSS | 2.3 (3.0) | 3.5 (4.3) | 3.8 (4.7) | 6.3 (5.7) |
| Barthel Index | 81.9 (22.1) | 76.2 (27.8) | 59.1 (35.9) | 55.6 (32.8) |
| Lesion Volume (mL) | 14.7 (25.5) | 27.4 (49.8) | 22.6 (37.2) | 25.6 (30.8) |
| Structural Disconnectivity (%) | 3.7 (4.1) | 5.5 (6.0) | 5.3 (5.2) | 6.7 (5.7) |

### Quantification of damage in NT-informed structural networks

In the Leipzig cohort, stroke lesions were delineated on clinical imaging data using the semi-automated Clusterize Toolbox, with manual editing. Oxford cohort lesions were manually delineated in MRIcron ^51,52^. For the normalization to Montreal Neurological Institute (MNI) standard space, the Clinical Toolbox was used in both cohorts. Please see Figure 1 for stroke lesion overlays. For details on image acquisition parameters, please refer to the original publications ^17,51^. To quantify lesion-induced damage within neurotransmitter (NT)-informed structural networks, binary lesion masks in MNI space were embedded into normative NT-weighted structural connectomes, as previously described^16,53,54^. Positron-emission tomography (PET)-derived receptor and transporter density maps covering 19 NT systems (including serotonergic, dopaminergic, cholinergic, glutamatergic, GABAergic, and noradrenergic systems) were obtained from Hansen et al. ^38^ and co-registered to a Human Connectome Project-derived normative connectome comprising 2 million streamlines^55^. Streamlines were weighted by the product of NT densities at their endpoints and normalized within each NT map to enable comparability across systems. Lesion masks were then overlaid onto these NT-informed connectomes, and NT-related network damage was quantified as the proportional summed weight of streamlines intersecting the lesion, yielding values between 0 (no damage) and 1 (complete damage). In addition, the proportion of disconnected unbiased streamlines was computed as a measure of overall structural network damage. This analysis is publicly available at https://github.com/phjkoch/NTDisconn.

**Figure 1.**
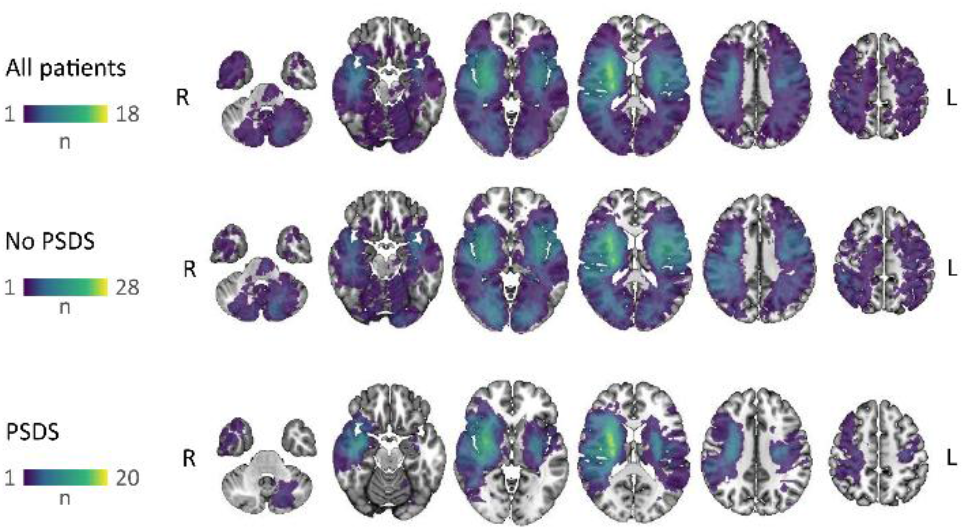
Lesion heatmap. Lesion distribution of patients in the combined cohort, displayed in standard MNI space. Colors indicate the number of patients with a lesion at each voxel. **Abbreviations**: PSDS: post-stroke depressive symptoms; R: right; L: left

### Statistical analysis

To assess the association between NT-related network damage and post-stroke depressive symptoms, we applied a two-step modeling framework designed for mechanistic inference rather than individual-level prediction. This framework combined partial least squares (PLS) regression with subsequent logistic regression analyses and was implemented in R (version 4.4.1). PLS regression was used to identify informative predictors while specifically addressing multicollinearity among NT-related network damage measures and the raw structural network damage (SL), which is unbiased with respect to any NT system. Predictors with Variable Importance in Projection (VIP) > 1 were included in further inferential outcome modeling. Although 10-fold cross-validation indicated that a single PLS component typically minimized prediction error, three latent components were retained for VIP score computation to improve the stability of variable weights and to capture distributed covariance patterns in multivariate data ^56-59^. Associations with PSDS were evaluated using generalized linear models with a binomial link function. All models were adjusted for age, sex, lesion volume, and stroke-related disability (BI and NIHSS). Predictor-level statistical significance was assessed using Wald-type z-tests.

Model fit was evaluated by comparing Akaike Information Criterion (AIC) values relative to models with covariables only, with a ΔAIC > 2 indicating meaningful model improvement ^20^. Finally, results were compared across cohorts to identify significant NT-PSDS associations that were consistent and independent of demographic covariates and lesion size.

## Results

Demographic and clinical characteristics stratified by PSDS status are summarized in Table 1. In both cohorts, patients with and without PSDS were comparable in age and sex distribution. PSDS was consistently associated with greater neurological symptom burden and functional impairment. Lesion volume and overall structural disconnectivity (SL) tended to be higher in patients with PSDS across cohorts, although, from a qualitative observational perspective, the magnitude of these differences varied between datasets. These cohort-specific profiles reflect differences in clinical composition and stroke characteristics, while providing complementary samples for examining convergent neurochemical correlates of PSDS.

Across both cohorts, lesion-induced network damage affected all investigated NT-related networks (Figure 2A). While the overall magnitude of NT-related network damage differed between cohorts, the relative involvement of NT systems showed qualitatively similar patterns. Partial least squares regression was applied to identify NT-informed network damage measures most strongly associated with the presence of PSDS. In the Leipzig cohort, VIP scores > 1 indicated that network disconnection related to serotonergic (5-HTT, 5-HT_2a_, 5-HT_4_), dopaminergic (D1), cholinergic (VAChT), and GABAergic systems contributed most strongly to the multivariate model (Figure 2B). In the Oxford cohort, a partially overlapping profile emerged, with GABA-, 5-HTT-, D1-, and VAChT-related network damage again, alongside the additional involvement of 5-HT_1a_ and M1. Of note, the unbiased global structural disconnection (SL) did not reach the threshold in either cohort.

**Figure 2.**
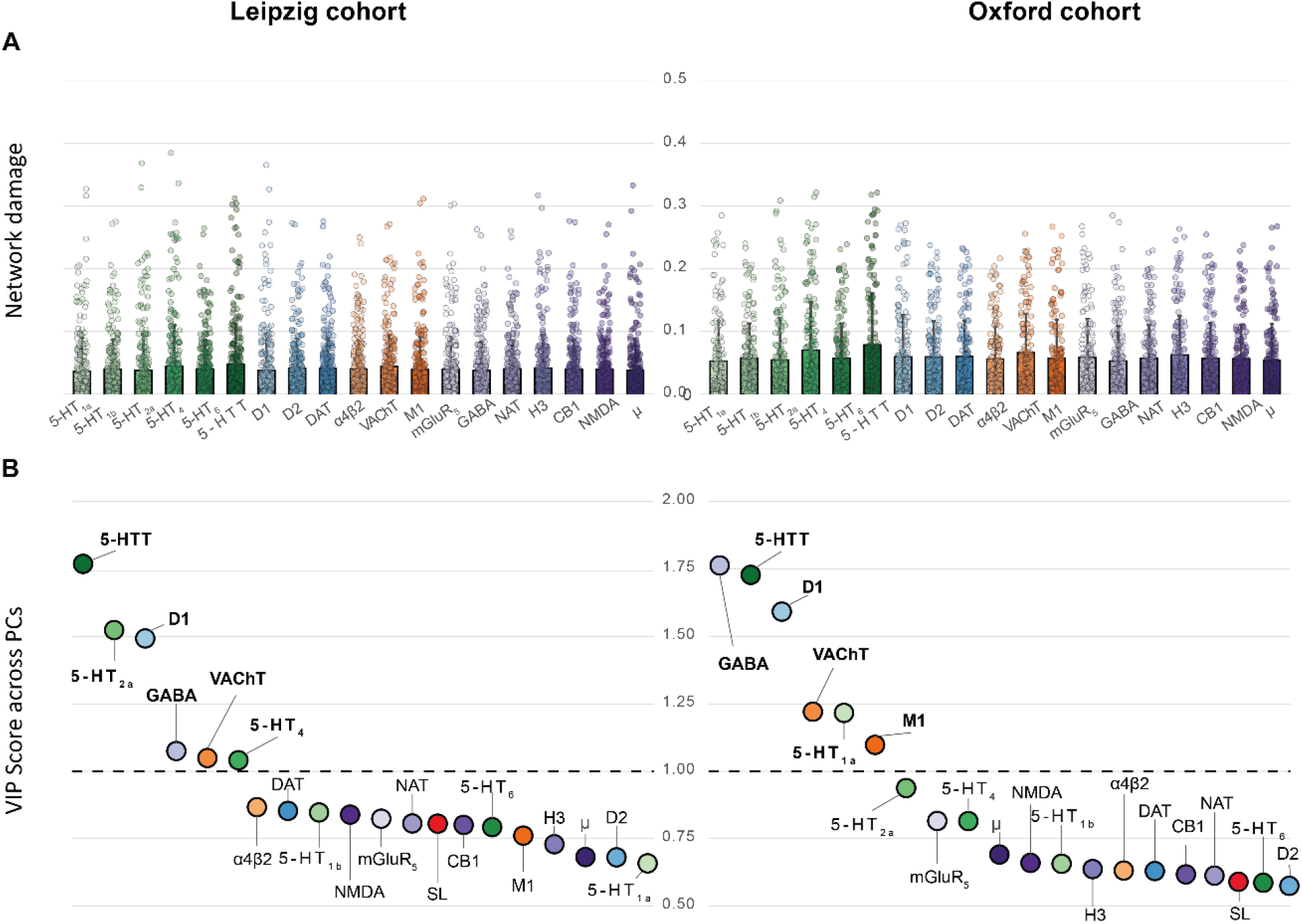
Neurotransmitter-informed network damage and PLS-based predictor selection. **A** Network damage scores (0-1) are plotted for the 19 neurotransmitter receptor and transporter networks for the two cohorts. **B** Informative predictors for inference of the presence of post-stroke depressive symptoms (PSDS) were selected based on partial least squares (PLS) regression and computation of cumulative variable importance in projection (VIP) scores across three latent components for stability of variable weights and to capture distributed covariance patterns in multivariate data. Informative predictors were defined as VIP>1. For visualization, data were grouped according to the NT system in green for serotonin, blue for dopamine, orange for acetylcholine, and purple for others. The global network damage (SL) is depicted as red. Predictors with VIP > 1 in were subjected to further GLM analysis for interpretation in the respective cohorts. **Abbreviations:** 5-HT_1a_, 5-HT_1b_, 5-HT_2a_, 5-HT_4_, and 5-HT_6_ denote serotonin receptor subtypes 1A, 1B, 2A, 4, and 6, respectively, while 5-HTT refers to the serotonin transporter. D1 and D2 indicate dopamine D1 and D2 receptors, and DAT the dopamine transporter. NAT denotes the norepinephrine transporter (noradrenaline transporter). VAChT denotes the vesicular acetylcholine transporter, M1 the muscarinic acetylcholine receptor M1, and α4β2 the α4β2 nicotinic acetylcholine receptor. mGluR_5_ indicates the metabotropic glutamate receptor subtype 5, NMDA the N-methyl-D-aspartate glutamate receptor, and GABA the γ-aminobutyric acid type A receptor. CB1 refers to the cannabinoid receptor type 1, µ to the μ-opioid receptor, and H3 to the histamine H3 receptor. SL denotes the structural lesion-induced disconnection score, quantifying the extent of white-matter disconnection caused by the stroke lesion.

NT-informed candidate networks identified by PLS were entered into covariate-adjusted generalized linear models to evaluate their association with PSDS (Table 2). In the Leipzig cohort, more severe damage to networks related to the serotonin transporter (5-HTT), to the vesicular acetylcholine transporter (VAChT), and to D1 was associated with increased odds of PSDS and led to meaningful improvements in model fit compared with covariate-only models. In contrast, associations with GABAergic network damage did not reach statistical significance after adjustment. In the Oxford cohort, higher 5-HTT- and VAChT-related network damage was also independently associated with PSDS and improved model fit, replicating the serotonergic and cholinergic effects observed in the Leipzig cohort.

**Table 2.**
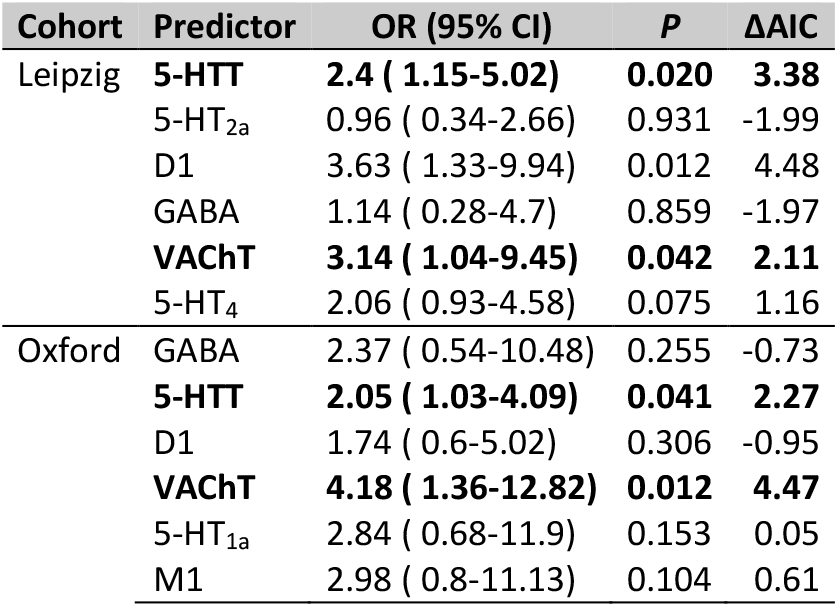
Association between neurotransmitter-informed network damage and post-stroke depressive symptoms (PSDS). Generalized linear models examine the association between raw NT damage scores and the presence of PSDS, defined as a HADS-D depression score above 7 at follow-up (yes/no, binary). Models are adjusted for age, sex, lesion volume, and neurological symptom burden operationalized by NIHSS and the Barthel index. Statistical significance is reported for each predictor within each model. Odds ratios (OR, for presence of PSDS) are given with 95% confidence intervals with a reference of 10% increase in NT/SL damage. Bolded neurotransmitters indicate convergent results across cohorts for each outcome. Model fit was evaluated by comparing Akaike Information Criterion (AIC) values with those of models with covariables only; a ΔAIC > 2 indicated meaningful model improvement.

However, the dopaminergic D1-related network involvement did not show a robust association in the multivariable analysis in the Oxford cohort. Taken together, these results indicate that serotonergic and cholinergic network damage, specifically involving the transporter of both systems, represents the most robust and reproducible neurochemical correlate of PSDS across independent datasets, whereas effects related to other NT systems appear cohort-specific and less stable.

## Discussion

The present findings suggest that post-stroke depressive symptoms are linked to stroke-related disruption of white matter pathways embedded within serotonergic and cholinergic signaling architectures. This perspective adds a novel molecular dimension to network-based accounts of PSDS by illustrating that PSDS are modulated by interactions between lesion topology and the neurochemical organization of large-scale brain networks.

### Serotonergic and cholinergic network disruption is the most robust and generalizable substrate

Across two independent cohorts, 5-HTT- and VAChT-related network damage was significantly associated with PSDS and improved model fit beyond clinical covariates, even after adjustment for age, sex, lesion volume, and neurological symptom burden. This convergent result provides a neurochemical anchor for PSDS vulnerability at the network level. Mechanistically, this observation integrates well with established evidence for serotonergic involvement in depression. Meta-analytic PET and post-mortem evidence suggest altered 5-HTT binding in major depression ^60^, and PET work indicates that binding differences are shaped by the serotonin system’s anatomy and regional expression patterns ^21,22^. Genetic studies further support serotonergic vulnerability in PSDS: meta-analyses demonstrate that functional 5-HTTLPR variants, particularly the short allele, increase susceptibility to post-stroke depression, whereas the long allele appears protective ^23,24^. From a network perspective, lesions intersecting brain-wide pathways embedded within serotonergic architectures may precipitate downstream reductions in serotonergic input to cortical and subcortical regions involved in affect regulation, motivation, and emotional control. This mechanism is consistent with circuit models of depression and with the fact that selective serotonin reuptake inhibitors exert their primary therapeutic effects via blockade of the serotonin transporter ^25,26^. However, large randomized controlled trials have shown that although fluoxetine modestly reduces the incidence of PSDS, it does not improve functional recovery and is associated with increased adverse events, including fractures and hyponatremia ^11,13,27^. These findings underscore the limitations of non-specific monoaminergic modulation and emphasize the need for targeted, biomarker-driven interventions. Beyond serotonergic networks, cholinergic network damage, as indexed by VAChT, was significantly associated with PSDS. Cholinergic neurotransmission, particularly via nicotinic acetylcholine receptors (nAChRs), has been implicated in mood regulation, cognition, and neuroimmune control. The α7-nAChR subtype, enriched in the hippocampus and prefrontal cortex, modulates monoaminergic release, synaptic plasticity, and microglial activation; engagement of α7-nAChR may exert antidepressant-like effects ^28,29^. Clinically, reduced availability of β2-containing nAChRs has been observed in major depression and correlates with illness burden ^30^, while pharmacological modulation of nAChRs has shown antidepressant potential in selected contexts ^31,32^. Importantly, VAChT regulates acetylcholine packaging and release; reduced VAChT expression can yield antidepressant-like behaviors in animal models and modulate serotonin and dopamine levels, suggesting cross-talk between cholinergic and monoaminergic systems ^33^. In PSDS, disruption of cholinergic pathways could therefore amplify vulnerability, particularly when depression co-occurs with cognitive slowing or apathy, which are linked to cholinergic dysfunction ^61,62^. In line with this, cholinergic involvement may represent an additional pathway rather than a primary marker.

### Dopaminergic network involvement as secondary, context-dependent signals

Dopaminergic D1-related network damage showed a significant association in Leipzig but not in the Oxford cohort, consistent with a cohort-dependent contribution. D1 receptors are key regulators of reward processing, motivation, goal-directed behavior, and cognitive control ^63,64^. Experimental and systems-level work indicate that D1 receptor signaling supports information flow through striatal pathways and can shape motivational and psychomotor components ^65,66^ that are often prominent in depressive syndromes. The cohort-specific nature of the D1 signal may reflect differences in lesion distribution, functional impairment, or the composition of depressive phenotypes (e.g., anhedonia vs. affective distress), underscoring that dopaminergic contributions to PSDS may be heterogeneous across clinical samples. Taken together, the lack of cross-cohort stability for D1-related network effects indicates that these findings should be interpreted as hypothesis-generating signals, highlighting potentially relevant but non-primary pathways that may refine mechanistic understanding and guide future, targeted investigations rather than serving as robust, cohort-independent markers of PSDS.

### Integrating lesion-network frameworks with neurochemical vulnerability

Our findings also help integrate lesion-network mapping insights with a molecular vulnerability framework. Lesion location alone has shown links to depression, with early studies reporting associations while also noting substantial dispersion of lesion sites among depressed patients ^34,43,46^. Lesion network mapping addresses this by showing that heterogeneous lesions may converge on a common functional circuit related to depression and by operationalizing “network damage” as a predictor of depressive outcomes ^7-9,34^. However, both lesion-based and network-based approaches leave a substantial proportion of inter-subject variability in PSD unexplained. Against this backdrop, our NT-informed approach suggests that neurochemical architectures, particularly serotonergic systems, might provide complementary information that extends established neuroimaging frameworks beyond topography alone, thereby refining the characterization of individual vulnerability to PSDS rather than replacing lesion-network models.

### Integration with the vascular depression framework

The present results also extend the concept of vascular depression, which posits that late-life depression arises from cumulative cerebrovascular injury disrupting frontal-subcortical circuits ^35,36^. Our data suggest that depressive symptoms may depend not only on the burden of white matter injury but also on whether lesions preferentially affect networks embedded within specific neurochemical architectures, most notably serotonergic pathways. In this framework, PSDS may emerge once a critical threshold of neurochemically relevant network damage is exceeded, consistent with cumulative models of vascular depression and reserve-related vulnerability ^37^.

### Limitations and future directions

Several limitations merit consideration. First, a limitation of the current approach is that NT-informed networks were derived from normative PET receptor atlases ^38^ and population-averaged structural connectomes, rather than from patient-specific molecular imaging or individual connectivity estimates. While normative connectomes are standard in lesion network mapping, they can capture general connectivity profiles ^39^ and may be advantageous relative to lower-quality single-subject diffusion MRI, especially when constructed from high-quality healthy-control datasets ^40^, they inherently lack the ability to reflect interindividual differences in receptor expression or network organization, potentially attenuating subject-level specificity in prediction models. Second, depressive symptoms were assessed at a single follow-up time point, limiting inferences regarding symptom trajectories, temporal precedence, or dynamic interactions between evolving network disconnection and affective outcomes after stroke. Third, although the present framework captures structural disconnection within neurochemically defined networks, it cannot directly probe molecular fingerprints or dynamic biological mechanisms such as neuroinflammation, epigenetic regulation, or treatment-mediated adaptations. Moreover, detailed information on antidepressant treatment, including timing, dosage, and treatment duration, was not available, limiting the ability to disentangle disease-related network vulnerability from potential medication-related effects. Accordingly, the observed associations should be interpreted as systems-level correlates rather than direct markers of underlying molecular processes. Importantly, despite adjustment for age, sex, lesion volume, and neurological deficit, the analytical approach remains fundamentally correlative and does not imply causal mechanisms linking serotonergic network disruption to PSDS. Fourth, no out-of-sample prediction was performed. While inferential analyses identified serotonergic network vulnerability as a consistent systems-level correlate of PSDS, the absence of cross- or externally validated predictions limits conclusions regarding individual-level prognostic utility; thus, the findings should be regarded as hypothesis-generating rather than clinically predictive. Finally, collinearity across NT systems remains a critical methodological consideration. To mitigate this, we employed PLS-based VIP measures and model the multivariate association pattern. These complementary strategies converged on serotonergic networks as the most stable contributors, whereas cholinergic and dopaminergic networks showed less consistent evidence, without excluding their potential relevance under specific clinical, temporal, or biological conditions.

Future longitudinal and multimodal studies integrating patient-specific connectivity, molecular imaging, inflammatory biomarkers, and genetic or epigenetic markers of serotonergic, cholinergic, and dopaminergic function are needed to determine whether serotonergic network disruption predicts PSDS trajectories, moderates treatment response, or delineates clinical contexts in which cholinergic or dopaminergic vulnerability constitutes an additional meaningful pathway. Prospective designs with repeated symptom assessments and independent validation cohorts will be essential to establish temporal directionality, improve generalizability, and clarify the translational potential of NT-informed network vulnerability in personalized post-stroke neuropsychiatric risk profiling.

## Data Availability

Anonymized behavioral and lesion data are available from the corresponding authors of the original papers on reasonable request.

## Acknowledgments

The authors acknowledge the use of artificial intelligence–based tools to support this work (ChatGPT version 5.2, OpenAI, San Francisco, CA, USA; and OpenEvidence, OpenEvidence Inc.). AI assistance was employed for (i) code troubleshooting for statistical analyses and data visualization, (ii) targeted literature search and synthesis, and (iii) language editing and structural refinement of the manuscript draft. All analyses, interpretations, and final manuscript content were critically reviewed, verified, and approved by the authors, who take full responsibility for the accuracy and integrity of the work.

## Author contributions

CRediT: Conceptualization: BMF, JK, MJM, ND, DS, RS; Data curation: JK, MJM, ND, DS; Formal Analysis: BMF, RS; Funding acquisition: RS; Investigation: ; Methodology: BMF, PJK, JF, TH, RS; Project administration: BMF, RS; Resources: ; Software: ; Supervision: RS; Validation: BMF; Visualization: BMF, JF, TH, RS; Writing – original draft: BMF, RS; Writing – review & editing: JK, MJM, PJK, JF, TH, GT, TM, FQ, ND, DS comparisons to prioritize neurochemical networks contributing most consistently to

